# Choroid plexus enlargement is associated with neuroinflammation and reduction of blood brain barrier permeability in depression

**DOI:** 10.1101/2021.09.30.21264226

**Authors:** Noha Althubaity, Julia Schubert, Daniel Martins, Tayyabah Yousaf, Maria A. Nettis, Valeria Mondelli, Carmine Pariante, Neil A. Harrison, Edward T. Bullmore, Danai Dima, Federico E. Turkheimer, Mattia Veronese, the NIMA Consortium

**Affiliations:** Department of Neuroimaging, Institute of Psychiatry, Psychology and Neuroscience, King’s College London, London, UK; Department of Radiological Sciences, College of Applied Medical Science, King Saud bin Abdulaziz University for Health Sciences, Riyadh, SA; Department of Psychological Medicine, Institute of Psychiatry, Psychology and Neuroscience, King’s College London, London, UK; Cardiff University Brain Research Imaging Centre (CUBRIC), Cardiff University, Cardiff, UK; Department of Neuroscience, Brighton and Sussex Medical School, University of Sussex, UK; Department of Psychiatry, School of Clinical Medicine, University of Cambridge, UK; Cambridgeshire and Peterborough NHS Foundation Trust, Cambridge, UK; Immuno-Psychiatry, Immuno-Inflammation Therapeutic Area Unit, GlaxoSmithKline R&D, Stevenage, UK; Department of Psychology, School of Arts and Social Sciences, City University of London, London, UK; Department of Information Engineering, University of Padua, Padua, Italy

**Keywords:** Choroid Plexus, Neuroinflammation, Depression, Blood Brain Barrier

## Abstract

**Background:** Recent studies have shown that choroid plexuses (CP) may be involved in the neuro-immune axes, playing a role in the interaction between the central and peripheral inflammation. Here we aimed to investigate CP volume alterations in depression and their associations with inflammation.

**Methods:** 51 depressed participants (HDRS score >13) and 25 age- and sex-matched healthy controls (HCs) from the Wellcome Trust NIMA consortium were re-analysed for the study. All the participants underwent full peripheral cytokine profiling and simultaneous [11C]PK11195 PET/structural MRI imaging for measuring neuroinflammation and CP volume respectively.

**Results:** We found a significantly greater CP volume in depressed subjects compared to HCs (t_(76)_ = +2.17) that was positively correlated with [^11^C]PK11195 PET binding in the anterior cingulate cortex (r=0.28, p=0.02), prefrontal cortex (r=0.24, p=0.04), and insular cortex (r=0.24, p=0.04), but not with the peripheral inflammatory markers: CRP levels (r=0.07, p=0.53), IL-6 (r=-0.08, p=0.61), and TNF-α (r=-0.06, p=0.70). The CP volume correlated with the [^11^C]PK11195 PET binding in CP (r=0.34, p=0.005). Integration of transcriptomic data from the Allen Human Brain Atlas with the brain map depicting the correlations between CP volume and PET imaging found significant gene enrichment for several pathways involved in neuroinflammatory response.

**Conclusion:** This result supports the hypothesis that changes in brain barriers may cause reduction in solute exchanges between blood and CSF, disturbing the brain homeostasis and ultimately contributing to inflammation in depression. Given that CP anomalies have been recently detected in other brain disorders, these results may not be specific to depression and might extend to other conditions with a peripheral inflammatory component.

## INTRODUCTION

Major Depressive Disorder (MDD) is a neuropsychiatric disorder associated with significant psychosocial impairment, recognized by the WHO as the leading cause of disability worldwide (Petralia, Mazzon et al. 2020). MDD is associated with mood changes such as sadness, crying, irritability, and anhedonia as well as psychophysiological symptoms such as insomnia, slowness of speech and action, loss of appetite, constipation, diminished sexual desire, and suicidal thoughts (Belmaker and Agam, 2008). Available antidepressant medications, which largely target monoamine pathways, are effective; however, more than 30% of depressed patients fail to achieve remission despite multiple treatment trials (Nettis et al., 2021). Treatment resistance in MDD has been associated with heighted peripheral immunity (Bekhbat et al., 2018).

MDD patients who experienced treatment resistance exhibit most of the cardinal features of inflammation, including elevations in inflammatory cytokines, acute phase proteins, chemokines, adhesion molecules, and inflammatory mediators such as prostaglandins in peripheral blood and CSF (Miller et al., 2009). Different studies have shown significant associations between inflammatory cytokines, in particular IL-1, TNF-α, and IL-6 that are markers of innate immune response, and depressive symptoms (Miller and Raison, 2016, Enache et al., 2019). Evidence of direct relationship between peripheral heightened immunity and MDD is supplied by the reproducible observation that the acute and chronic administration of cytokines (or cytokine inducers such as lipopolysaccharide (LPS) or vaccination) can cause behavioural symptoms that overlap with those found in MDD (Miller and Raison, 2016). For example, 20% to 50% of patients receiving chronic IFN-alpha therapy for the treatment of infectious diseases or cancer develop MDD (Lotrich, 2009). Moreover, inducing acute IFN-alpha in healthy subjects results in the elevation of peripheral inflammation immune markers such as C-reactive protein, TNF-alpha, and IL-6 and is accompanied with depressive symptoms (Nettis et al., 2020).

The mechanisms linking peripheral immunity to changes in central nervous system and mood changes in MDD are still under investigation. The hypothesis of a potential direct action of peripheral cytokines trespassing the blood brain barrier (BBB) to activate brain microglia (D’Mello et al., 2009) has spurred a number of imaging studies of neuroinflammation in the central nervous system (CNS) in MDD that have been conducted using positron emission tomography (PET) and radiotracers targeting the 18kDa mitochondrial translocator protein (TSPO), a protein that is consistently upregulated in activated microglia (see review in (Mondelli et al., 2017)). These have revealed mild microglial activity in depressed subjects (Schubert et al., 2021b); however, evidence of a relationship between central and peripheral inflammation for PET imaging has been lacking (Schubert et al., 2021b). Instead, there is very recent evidence of a relationship between heightened peripheral immunity and the reduction of the brain barriers permeability, both for the BBB and blood-CSF barrier (BCSFB) at the choroid plexus (CP) (Turkheimer et al., 2020).

This has led us to hypothesize a different model of peripheral-to-central inflammatory interaction whereas brain barriers react to increases in circulating inflammatory messengers by reducing peripheral-to-central solute transfer (Turkheimer et al., 2020). This may be protective in the short term but, if the inflammatory status becomes chronic, may disturb brain homeostasis and induce microglia to react to the disturbed environment. Very recent work on immune peripheral activation via LPS in mice and rats supports this view by showing that sickness behaviour is not mediated by microglia, hence their activation is a secondary result of the peripheral immune challenge (Vichaya et al., 2020).

The model above leads to a novel focus on a particular brain structure, the CP, that is an important part of the brain barriers, plays an active role in immune responses in the nervous system (Meeker et al., 2012) and, importantly, can be measured in-vivo using MRI imaging: the choroid plexus **(Figure 1)**.

**Figure 1:**
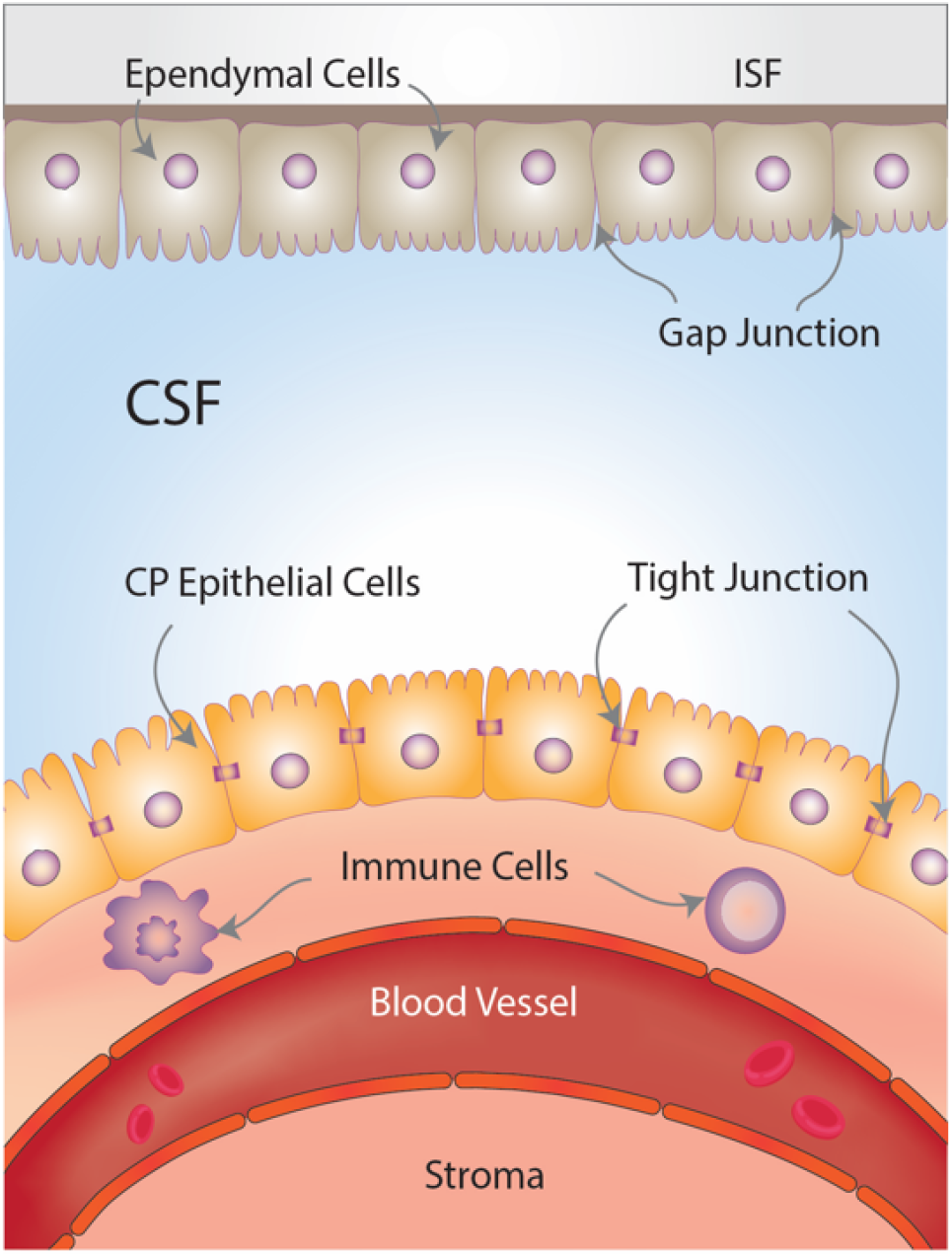
Choroid plexus (CP) and brain-CSF-barriers (BCSFB). The CP is located at the base of each of the four brain ventricles and is composed of epithelial cells surrounded by connective stroma and blood vasculature, which forms the BCSFB. The epithelial cells are connected by tight junctions in the apical border which face the cerebrospinal fluid (CSF) and are covered by microvilli, while the basolateral border faces the blood vasculature. The ependymal cells cover the roof of the ventricles and are connected by gap junctions, which facilitate the exchange of electrolytes and some solutes between the CSF and the interstitial fluid (ISF).

The primary role of the CP is to produce the cerebrospinal fluid (CSF) and form the BCSFB (Liddelow, 2015). The CP is an epithelial-endothelial convolute, consisting of a highly vascularised stroma with connective tissue and epithelial cells and is located in the basal lamina of the four brain ventricles (Shen, 2018, Brown et al., 2004, Khasawneh et al., 2018). It is responsible for producing 75% of CSF, and through the action of active transporters and channels mediates the movement of water and solutes across the epithelium (Ghersi-Egea et al., 2018b). CP epithelial cells are connected via tight junctions, which play a fundamental role in regulating the permeability and integrity of the BCSFB (Ghersi-Egea et al., 2018a). The fenestrations in the choroidal vessels are less restricted compared to cerebral vessels, and allow ions and small molecules such as amino acids to diffuse into the CP epithelial cells, which can then be actively transported actively into the CSF (Maharaj et al., 2008, Liddelow, 2015). The BCSFB together with the BBB control the traffic between the blood, the CSF, and the brain parenchyma (Saunders et al., 2012). There has been a recent increase of interest in the CP due to its important role in regulating brain homeostasis and, particularly, the neuroinflammatory response (Schwartz and Baruch, 2014). CP immune cells and intraventricular macrophages provide the first line of defence against brain infections and inflammation (Pollak et al., 2018, Yang et al., 2021). Prolonged inflammation may contribute to the infiltration of the CP by immune cells, and subsequently into the CSF and into the brain parenchyma (Marques and Sousa, 2015).

There is a growing body of literature that recognises the importance of CP morphology in psychiatric conditions. In subjects with schizophrenia, CP enlargement has been found to be associated with chronic stress (Zhou et al., 2020), a condition often associated with MDD and inflammation. A more direct correlation between CP volume and levels of peripheral IL-6 has been documented by Lizano and colleagues in psychosis (Lizano et al., 2019) whereas CP enlargement was also associated with reductions in gray matter and amygdala volume as well as ventricle enlargement and consequential deteriorated cognitive status (Lizano et al., 2019). Note that CP enlargement is also observed in patients with mild cognitive impairment and Alzheimer’s disease, for which MDD is a known prodrome (Tadayon et al., 2020). However, few studies suggest a potential direct link between CP structural and functional alterations and depression. Gene expression analysis from a cross-sectional post-mortem study has identified multiple differential mRNA expression in CP samples obtained from MDD patients (Turner et al., 2014, Devorak et al., 2015). ICAM-1 and VEGF transcripts are notable examples as they are known to play an important role in depression, being part of the tissue inflammatory response. They are also known to affect BBB permeability and in fact their peripheral measures serve as inflammatory markers (Müller, 2019, Clark-Raymond and Halaris, 2013).

In this study we wanted to test the model of peripheral-to-central inflammation sketched above by investigating for the first time the association between CP volume and neuroinflammation in a depressed cohort, using structural MRI and 18kDa translocator protein (TSPO) positron emission tomography (PET) data. We hypothesized that if the BCSFB were indeed a mediator between peripheral and central inflammation in depression, we would observe a direct correlation between central inflammation, here marked by TSPO parenchymal expression, and CP volume.

## Methods

### Dataset

Data for this study were collected from a network of clinical research sites in the United Kingdom as part of the Biomarkers in Depression Study (BIODEP, NIMA consortium, https://www.neuroimmunology.org.uk/) and included 51 depressed participants and 25 healthy controls (HCs). Depressed subjects who had taken anti-depressive treatment with Hamilton Depression Rating scale (HDRS)>13 and untreated subjects with HDRS>17 were included. In total, there were 9 untreated subject and 42 were medicated. An overview of demographic and clinical characteristic data is reported in **Table 1**, while full details of the dataset are outlined in the previous publication (Schubert et al., 2021b). Participant’s inclusion criteria were as follows: no history of other neurological disorder, no current alcohol and/or drug abuse, no contribution in any clinical drug trials during the previous year. Participants were not experiencing any other medical disorders or undergoing any treatment that could affect the accuracy of the study’s results. Pregnant or breastfeeding women were excluded. HC subjects were age- and sex-matched with the depressed subjects, and they did not have a history of clinical depression or antidepressant treatment. Ethical approval was obtained from the Institutional and/or National research committee and and with the 1964 Helsinki declaration and its later amendments or comparable ethical standards. All participants provided written informed consent prior to the study. The BIODEP study was approved by the NRES Committee East of England Cambridge Central (REC reference:15/EE/0092) and the UK Administration of Radioactive Substances Advisory Committee.

**Table 1.** Demographics and clinical characteristics for depressed subjects and healthy controls (HCs)

| **Variable** | **Depressed subjects**<br>**(n = 51)** | **Healthy controls**<br>**(n = 25)** | **p Value** |
| --- | --- | --- | --- |
| *<u>Demographics</u>* |  |  |  |
| Age, mean ± SD, years | 36.2 ± 7.3 | 37.3 ± 7.8 | 0.56 |
| Male, No. (%) | 15 (29) | 11 (44) | 0.21 |
| Weight, mean ± SD, kg | 80.3 ± 14.4 | 73.7 ± 15.1 | 0.10 |
| BMI, mean ± SD, kg/m<sup>2</sup> | 27.2 ± 4.0 | 24.2 ± 4.8 | <0.01* |
| Blood Variables, (MDD No.; HC No.),<br>mean ± SD: | | | |
| CRP mg/L (51; 25) | 2.9 ± 2.8 | 1.1 ± 0.9 | <0.01* |
| IL6 (35;12) | 1.1 ± 1.63 | 0.6 ± 0.4 | 0.35 |
| TNF-α (34;11) | 2.7± 0.9 | 2.3 ± 0.5 | 0.25 |
| IFN-γ (35;11) | 4.0 ± 0.7 | 4.2 ± 0.0 | 0.26 |
| IP-10 (49;22) | 402 ± 152 | 428 ± 101 | 0.46 |
| Lymphocytes (47;23) | 30.9 ± 7.2 | 30.2 ± 10.2 | 0.79 |
| Monocytes (47;23) | 6.8 ± 2.1 | 6.6 ± 2.5 | 0.78 |
| Neutrophil's total (47;23) | 58.8 ± 8.3 | 59.3 ± 11.3 | 0.80 |
| Neutrophils absolute (47;23) | 3.9 ± 1.5 | 4.1 ± 1.4 | 0.48 |
| Albumin (47;23) | 44.8 ± 2.7 | 45.3 ± 2.1 | 0.44 |
| Cholesterol (47;23) | 5.2 ± 1.2 | 4.9 ± 1.2 | 0.35 |
| VEGF (35;11) | 82 ± 55.5 | 136 ± 47.8 | <0.01* |
| Depressive Clinical Scores, (MDD No.),<br>mean ± SD: |  |  |  |
| HDRS (51) | 18.5 ± 3.7 | 0.6 ± 0.9 | <0.001* |
| Childhood Trauma score (50) | 54.3 ± 14 | 38.2 ± 5.4 | <0.001* |
| Perceived Stress score (48) | 26.7 ± 4.3 | 10 ± 5.9 | <0.001* |
| ICV, mean ± SD, mm<sup>3</sup> | 1,562,594 ± 137,333 | 1,550,724 ± 185,887 | 0.76 |

Abbreviations: NA, not applicable

### Clinical Assessments

For the clinical evaluation of depression, HCs and depressed subjects underwent a number of psychiatric assessments: the Hamilton Depression Rating scale (HDRS), the Standard Clinical Interview for DSM-5, the Beck Depression Inventory, the Spielberger State-Trait Anxiety Rating Scale, the Chalder Fatigue Scale, the Snaith-Hamilton Pleasure Scale, the Childhood Trauma score, and the Perceived Stress score. A venous blood sample was collected from all participants to assess peripheral blood parameters including interlukin-6 (IL-6), tumor necrosis factor-alpha (TNF-α), interferon gamma (IFN-γ), interferon gamma-induced protein 10 (IP-10), lymphocytes, monocytes, neutrophil’s total, neutrophils absolute, albumin, cholesterol, VEGF, and C-reactive protein (CRP) concentration. CRP was used as the main marker of a peripheral inflammation, consistent with other studies in MDD (Howren et al., 2009, Haapakoski et al., 2015).

### MRI and PET Imaging

For all the subjects, imaging data were obtained using a GE SIGNA simultenous PET/MR scanner (GE Healthcare, Waukesha, USA). All MRI acquisitions included a high resolution T1-weighted image for structural imaging. PET imaging consisted in a bolus injection of [^11^C]PK11195 (target dose ∼350 MBq, mean injected dose 361+ 53MBq) followed by a 60-minute dynamic acqusition. A multi-subject atlas method was used for attenuation correction and included improvement for the MRI brain coil component (Burgos et al., 2014). Additional corrections (including scatter, random, normalisation, and deadtime correction), were performed using the standard console software that applied PET/MR reconstruction algorithm correction. Additional information on the PET and MRI protocols can be found in the original report on these data (Schubert et al., 2021a).

### CP Volume and brain volume measurement

The CP of the lateral ventricles of all subjects was manually segmented on the axial, sagittal, and coronal planes of the high resolution T1-weighted image using the Analyze software (v.12, https://analyzedirect.com) **(Figure 2)**. The image intensity was adjusted to assist in the localization of the region of interest and its anatomical boundaries. The CP was traced slice by slice starting from the axial plane followed by coronal and sagittal planes. After the region contouring, the CP volume was calculated for each subject using the Analyze software. The SPM12 (http://www.fil.ion.ucl.ac.uk/spm) was used to perform tissue segmentation for gray matter, white matter, and CSF (Heinen et al., 2016). High resolution T1 weighted images were smoothed and normalised into MNI standard space using the DARTEL algorithm via the SPM12 for brain volume calculation (Ashburner, 2007). The total brain volume was obtained by the sum of gray and white matter volume (Heinen et al., 2016). Additionally, the Freesurfer software (v.6.0, http://surfer.nmr.mgh.harvard.edu/), was used to estimate the total intracranial volume (ICV) (Fischl et al., 2002, Jovicich et al., 2009), which was used as a covariate in the statistical analysis (Pintzka et al., 2015).

**Figure 2:**
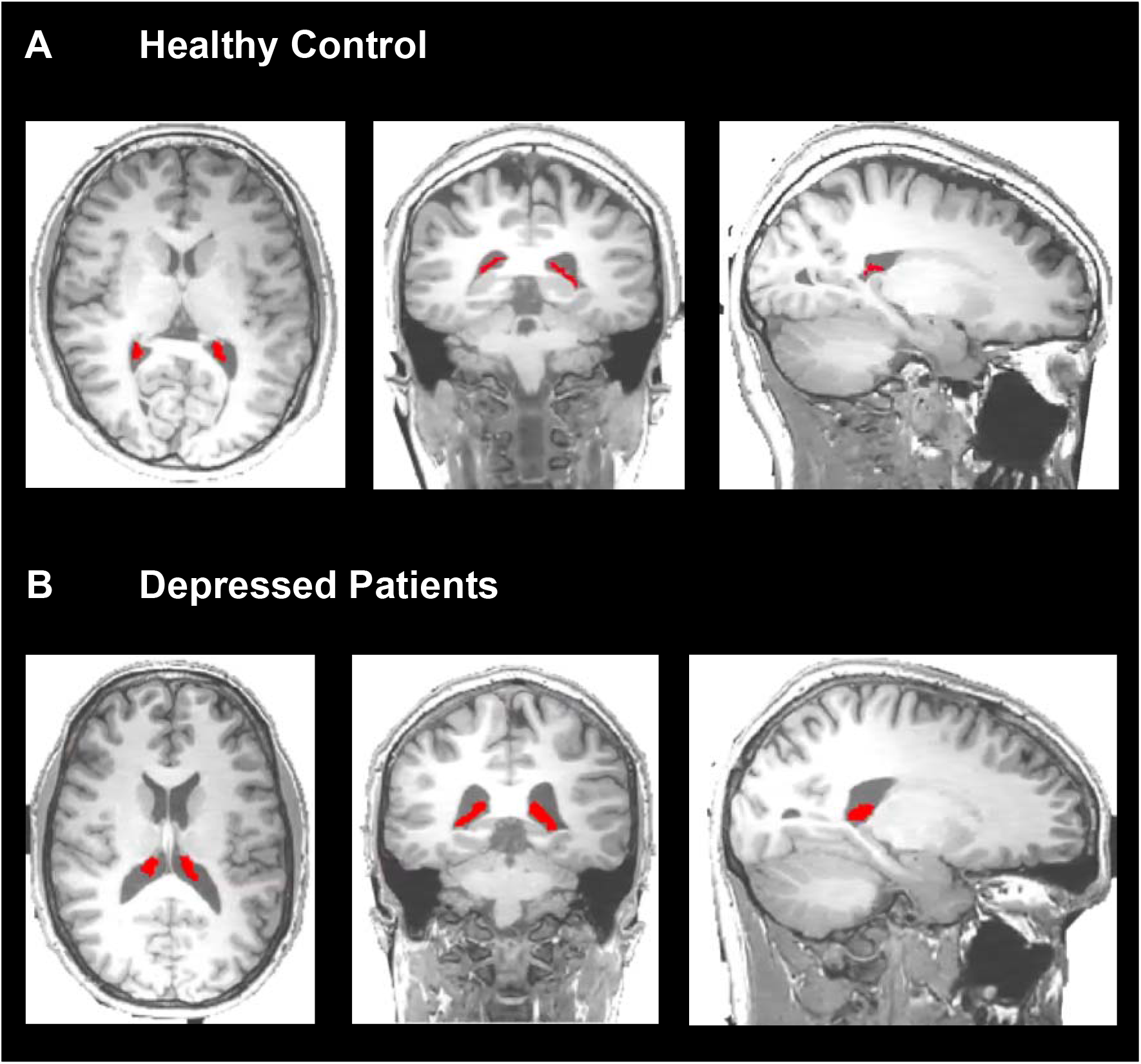
Choroid plexus (CP) segmentation in HCs and depressedsubject. T1-weighted MRI images with the manual segmentation of the left and right CP (red) on the axial, coronal, and sagittal planes for one representative control (A) and one depressed subject (B).

### PET Analysis

The pre-processing of the [^11^C]PK11195 PET imaging including frame-by-frame motion correction, PET-MRI alignment, brain masking, and atlas-based region extraction was performed using the MIAKAT software (www.miakat.org). Processed data were hence used for both the quantification of TSPO distribution in brain parenchyma (as a proxy of neuroinflammation) as well as to assess the tracer exchange between blood and CSF in the lateral ventricles. This second method had been previously validated and used to demonstrate CSF dynamic alterations in Alzheimer’s disease (De Leon et al., 2017, Schubert et al., 2019) and multiple sclerosis (Schubert et al., 2019). The lateral ventricles were manually segmented using T1-weighted structural MRI and ITK-SNAP software as was done in the original reference (Schubert et al., 2019, Acabchuk et al., 2015), and then eroded by two voxels in native space (5.2 mm) to reduce the effect of the partial volume (Yushkevich et al., 2006, Turkheimer et al., 2020). The [^11^C]PK11195 PET activity extracted from the eroded lateral ventricles was used to calculate the standardized uptake value ratio (SUVR) normalized by a reference region at 60 minutes, and the area-under-curve (AUC) which included activity of the lateral ventricles between 30 and 60 minutes (AUC_30-60_) (Turkheimer et al., 2020). For the quantification of TSPO, the simplified reference tissue model was used to calculate the distribution volume ratio (DVR) of [^11^C]PK11195 in brain (Turkheimer et al., 2007). The DVR estimates for the anterior cingulate cortex (ACC), prefrontal cortex (PFC), and insula (INS), were extracted to assess the level of brain inflammation as these regions serve important roles in mood regulation (Goldin et al., 2008), and the ACC has been previously shown to be involved in inflammation-associated mood deterioration (Harrison et al., 2009, Torres-Platas et al., 2014). The [^11^C]PK11195 DVR in CP was estimated from the parametric maps after performing partial volume correction using the Richardson-Lucy deconvolution method with 6mm point spread function provided by the PETPVC toolbox (Thomas et al., 2016). For both methods, the same reference region derived from a supervised clustering of dynamic PET images was applied (Schubert et al., 2021a). Additional details about the PET image processing and quantification are found in our previous studies (Turkheimer et al., 2020, Schubert et al., 2021b)

### Statistical Analysis

Statistical analyses were performed using SPSS software (version 25.0, Chicago, IL). The Shapiro-Wilk’s test was used to test for normality of the data. An independent sample t-test was used to examine the difference of CP volume between the HCs and depressed subjects. Partial correlations were used to analyse the relationship between CP volume and depressive clinical scores (HDRS, the Childhood Trauma, and Perceived Stress score), while covarying for ICV. Partial correlation tests were used to investigate the association between the CP volume and blood to CSF exchange measures (SUVR and AUC_30-60_) while covarying for the ICV and group. Partial correlation of CP volume measures and brain TSPO values (ACC, PFC, and INS) and brain volume were also investigated while covarying for possible confounding factors including ICV and group. The same analyses were repeated voxelwise with SPM12, using FWE for multiple comparison corrections.

### Imaging transcriptomics

We leveraged transcriptomic data from the Allen Human Brain Atlas (AHBA) (Hawrylycz et al., 2012) to explore possible associations between the brain map depicting the correlations between CP volume and TSPO and post-mortem brain gene expression. With this analysis, we aimed to gain further insight about potential biological pathways explaining regional vulnerability to spatial variability in the association between CP volume and brain TSPO across subjects.

We used the *abagen* toolbox (https://github.com/netneurolab/abagen) to process and map the transcriptomic data to 83 parcellated brain regions from the DK atlas (Desikan et al., 2006), as described in *Martins et al. (Martins et al., 2021)*. We applied the normalization for cortical and subcortical regions separately, as suggested by *Arnatkeviciute et al*., *2019* (Arnatkeviciute et al., 2019). After pre-processing, we retained regional gene expression data from 15,633 genes, corresponding to genes with expression higher than background noise. We then used partial least square regression (PLS) to investigate associations between the t-statistics quantifying the regional correlations between CP volume and TSPO and brain gene expression. We focused on both cortical and subcortical regions from the left hemisphere only. This choice was motivated by scarcity of data in the AHBA for the right hemisphere.

Partial least square regression ranks all genes by their multivariate spatial alignment with the regional strength of the association between CP volume and TSPO. The first PLS component (PLS_1_) is the linear combination of the weighted gene expression scores that have a brain expression map that covaries the most with the map of the association between CP volume and TSPO. We confirmed PLS_1_ explained the largest amount of variance by testing across a range of PLS components (between 1 and 15) and quantifying the relative variance explained by each component. The statistical significance of the variance explained by each PLS model was tested by permuting the response variables 1,000 times, while considering the spatial dependency of the data by using a spin test (Alexander-Bloch et al., 2013a, Alexander-Bloch et al., 2013b, Vasa et al., 2018). Since PLS_1_ alone explained the largest amount of variance in the imaging dependent variable, we focused our following analyses on this component.

The error in estimating each gene’s PLS_1_ weight was assessed by bootstrapping (resampling with replacement of the 34 brain regions), and the ratio of the weight of each gene to its bootstrap standard error was used to calculate the *Z* scores and, hence, rank the genes according to their contribution to PLS_1_ (Whitaker et al., 2016). Genes with large positive PLS_1_ weights correspond to genes that have higher than average expression in regions where the association between CP volume and TSPO is more strongly positive. Mid-rank PLS weights showed expression gradients that are weakly related to the pattern of the regional association between CP volume and TSPO.

We then used the list of genes ranked by the respective weights according to the PLS_1_ component to perform gene set enrichment analyses for biological pathways (gene ontology) and genes expressed in different brain cell types, as identified in a previous single-cell transcriptomic study of the human brain (Lake et al., 2018). We implemented these analyses using the GSEA method of interest of the Web-based gene set analysis toolkit (*WebGestalt*) (Zhang et al., 2005).

For comparative purposes, we used the same method to understand the spatial variability of changes in brain TSPO between subjects with depression and HCs.

## RESULTS

### Demographic and Clinical Characteristics

Subjects with depression and HCs were matched in terms of age (p=0.56) and sex (p=0.21), and no difference was shown in terms of the ICV between groups (p=0.76). BMI was significantly higher in depressed subjects compared to HCs (t_(76)_=3.52, p=0.001).

Peripheral blood parameters were all comparable between two groups (**Table 1**), with the exception of CRP and VEGF levels. Depressed subjects had higher blood CRP (t_(76)_ =3.14, p=0.002) and lower VEGF (t_(46)_ =-2.90, p=0.006) levels compared to HCs. While CRP differences were expected given the recruitment parameters (Schubert et al., 2021b), the difference in VEGF level was not.

In terms of clinical variables, the HDRS, Childhood Trauma scores, and the Perceived Stress score were significantly higher in depressed subjects compared to HCs (t_(76)_ =23.6, p<0.001), (t_(75)_ =5.5, p<0.001), (t_(73)_ =13.8, p<0.001) respectively **(Table 1)**.

### Depressed subjects have larger CPs compared to HCs

The CP volume was log10-transformed to induce normality in the data distribution as confirmed by Shapiro-Wilk’s test (W_(76)_ =0.98, p=0.23). The CP volume was greater in patients with depression compared to HCs (mean value of depressed patients:1710.29 ±408.80; mean value HCs: 1513.96±459.80; t_(76)_=2.17, p=0.03) (**Figure 3)**. In both groups, the CP volume was strongly correlated with ICV (r=0.25, p=0.03) and sex (F=4.7, p=0.03), but not with age (p=0.72) or BMI (p=0.19). However, when both ICV and sex were included as covariates, only ICV remained significant. After adjusting for ICV, the CP volume remained significantly higher in depressed subjects compared with HCs (F_(76)_=4.6, p=0.04) (**Figure 3)**.

**Figure 3:**
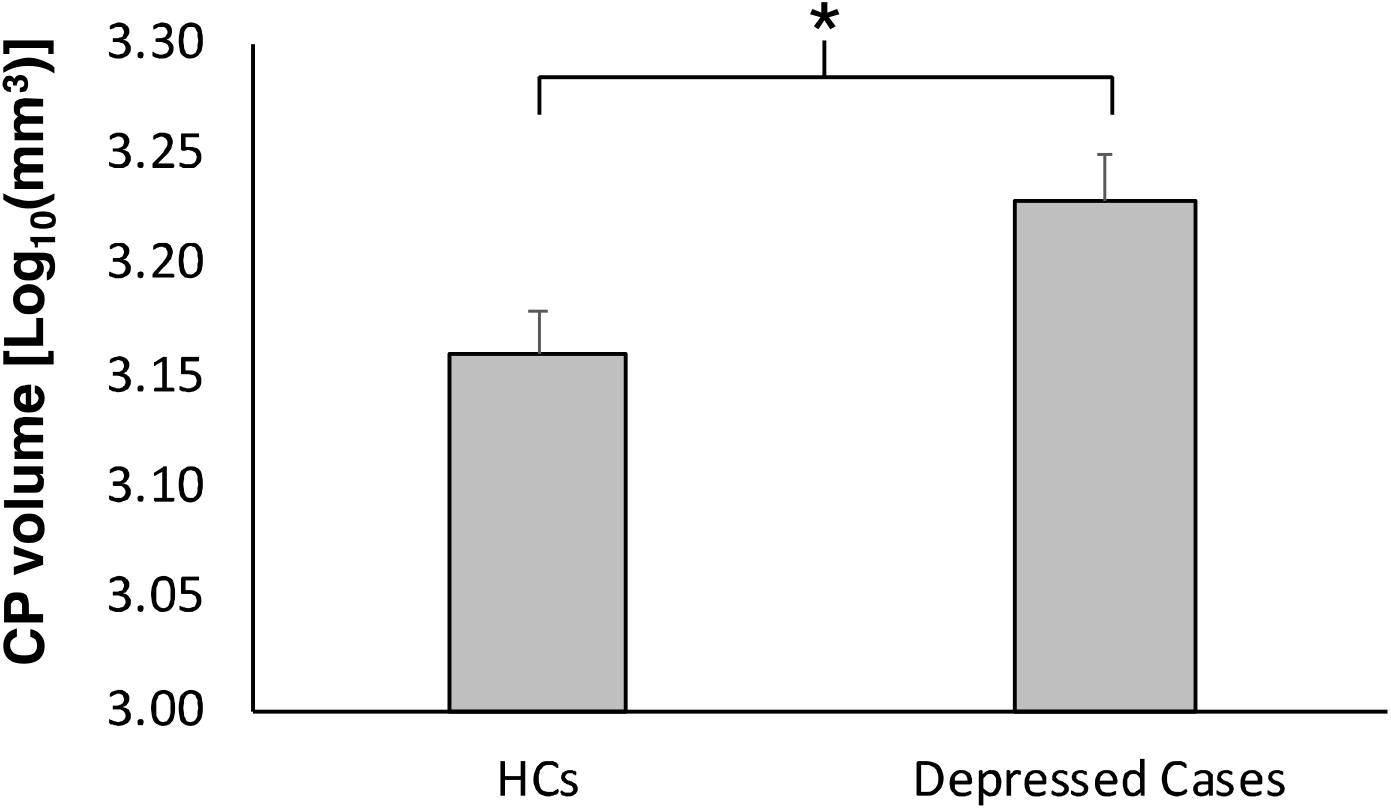
Choroid plexus (CP) volume in depressed subjects and healthy controls (HCs) Mean difference of CP volumes between HC and depressed groups (F=5.30, p=0.02). Error bars indicate standard error. The analysis is corrected for the intracranial volume.

After adjusting for ICV, the CP volume of depressed subjects showed a negative association with Perceived Stress score (r=-0.30, p=0.05); however, the association was not significant following multiple comparisons correction. No association was found with any other clinical variable.

### CP volume is not associated with peripheral inflammatory markers

When using the full sample, the CP volume was not associated with the CRP levels (r=0.07, p=0.53), IL-6 (r=-0.08, p=0.61), TNF-α (r=-0.06, p=0.70), or VEGF (r=0.12, p=0.39). Similarly, no association between CP volume and peripheral inflammatory markers was found when depressed and HC groups were analysed separately. However, an inverse association was found between the albumin level in plasma and CP volume (r=0.27, p=0.03) after adjusting the ICV, lateral ventricles volume and group.

### CP volume is positively associated with central inflammation and inversely with CSF clearance

Using the full dataset (N=76), CP volume measures were positively correlated with [^11^C]PK11195 DVR in the ACC (r=0.28, p=0.02), PFC (r=0.24, p=0.04) and INS (r=0.24, p=0.04) (**Figure 4)**. All these correlations were corrected for group and ICV, although only ICV was a significant covariate in the regression for ACC and INS (**Figure 4)**. When analysis was repeated in the depression cohort only, no correlation was detected between the CP volume and [^11^C]PK11195 DVR in any of the main ROIs (ACC: r=0.20, p=0.40; PFC: r=0.18, p=0.18; INS: r=0.17, p=0.2). No correlation was found between CP volumes and total cortical [^11^C]PK11195 DVR (r=-0.21, p=0.07) nor with whole brain [^11^C]PK11195 DVR (r=-0.05, p=0.56). CP volume and [^11^C]PK11195 DVR in CP were instead correlated both in the full sample (r=0.34, p=0.005) and in the depression cohort only (r=0.39, p=0.001), after correction for ICV, lateral ventricle volumes, and group.

**Figure 4:**
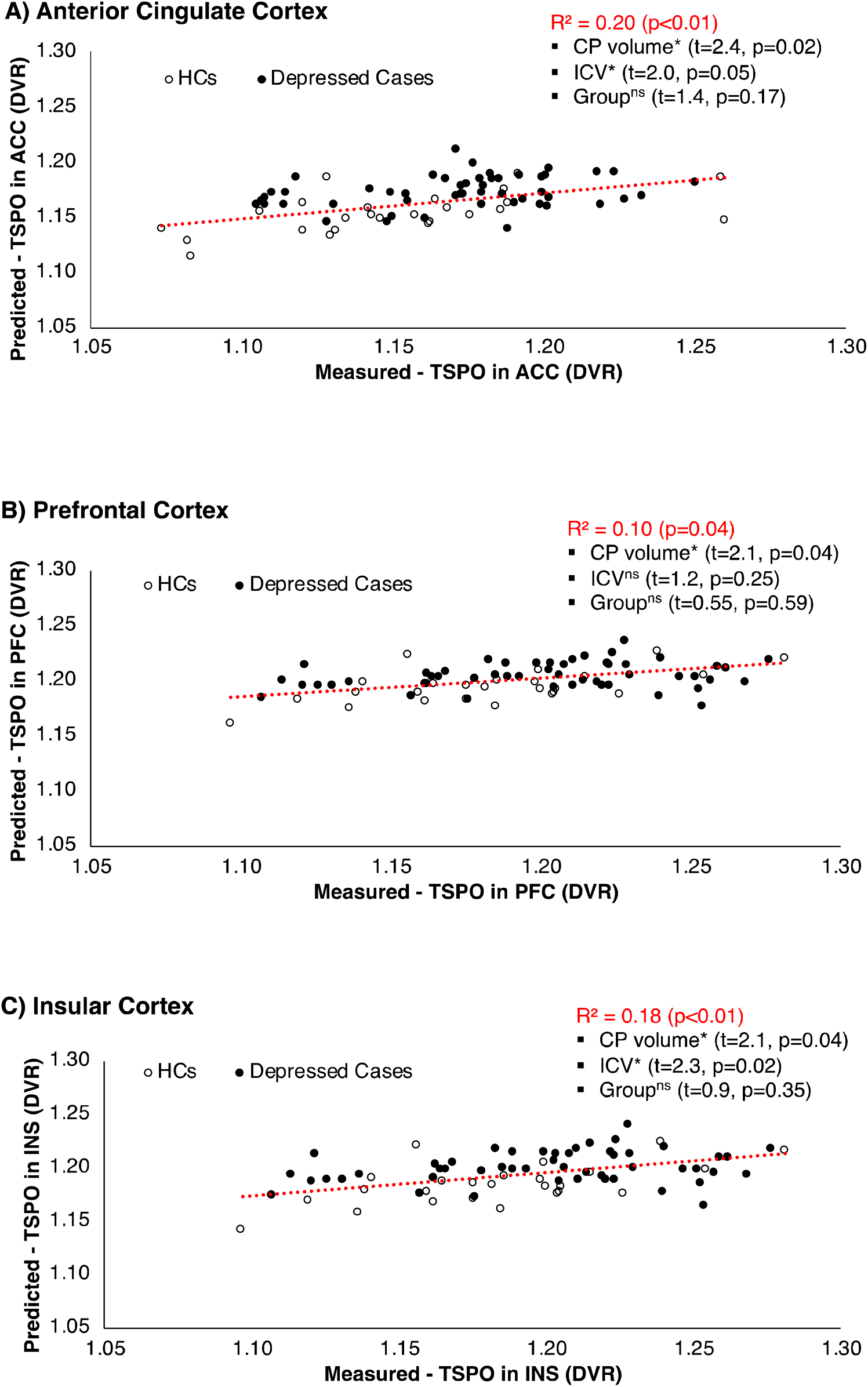
Correlation between the choroid plexus (CP) volume and brain inflammation in healthy controls (HCs) and depressed subjects in ACC (A), PFC (B) and INS (C). *indicates statistical significance (p-value<0.05). ^ns^ indicates non-significant results.

The CP volume exhibited a negative association with the blood-to-CSF radiotracer exchange as measured by the lateral ventricle SUVR (r=-0.23, p=0.05), following correction for group and ICV. A similar association was found for radiotracer AUC_30-60_ in lateral ventricle (r=-0.26, p=0.02) (**Figure 5)**. However, the depression cohort did not show a significant association between the CP volume and the SUVR (r=-0.19, p=0.19) nor AUC_30-60_ (r=-0.20, p=0.16).

**Figure 5:**
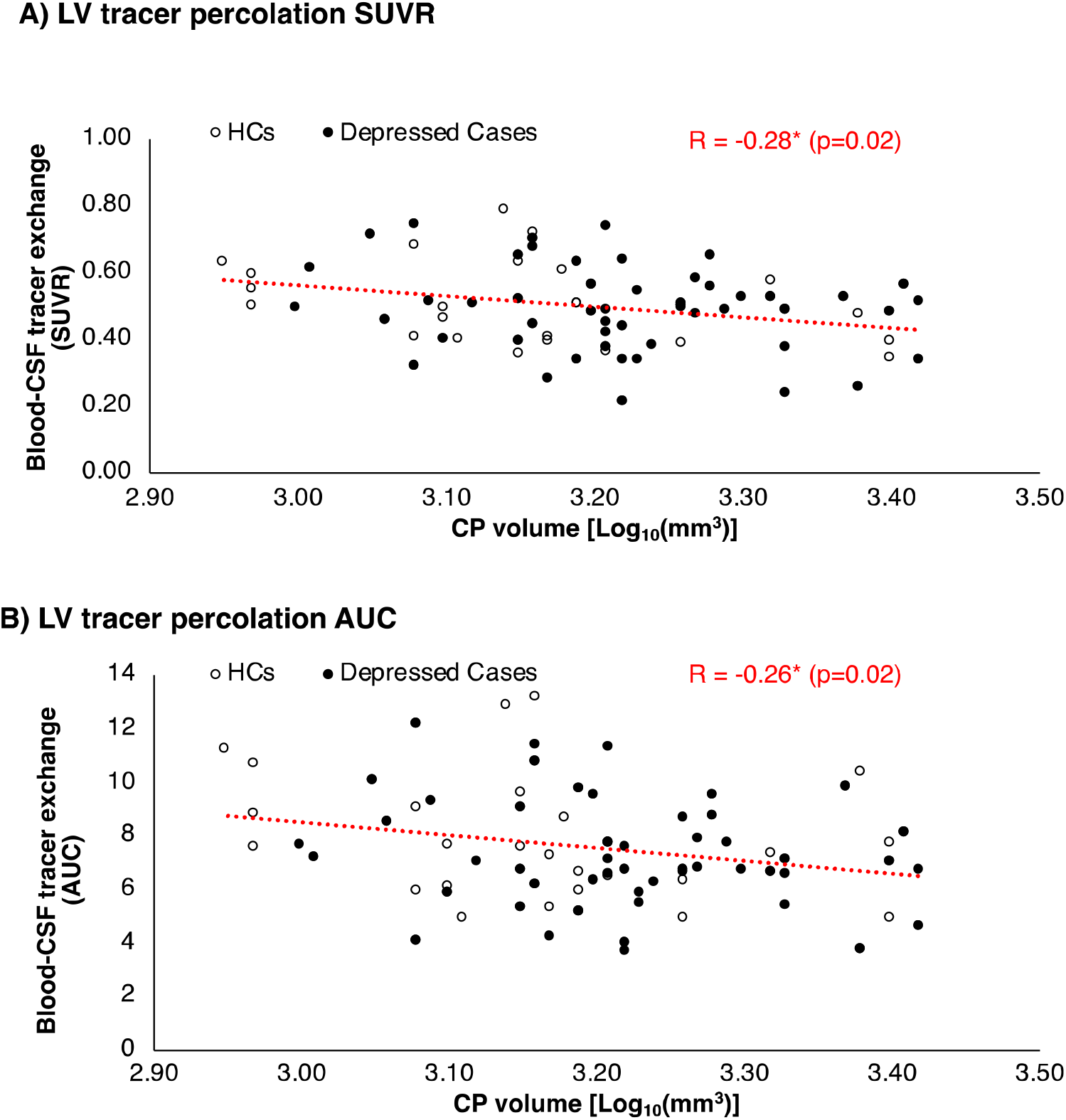
Inverse association between choroid plexus (CP) volume and CSF-blood tracer exchange measured by [^11^C]PK11195 PET uptake (SUVR and AUC, respectively) in lateral ventricles (LV). * indicates statistical significance (p-value<0.05).

### CP volume is not associated with brain volume

No association was detected between the CP volume and brain volume (r=0.16, p=0.17), nor with total grey matter volume (r=0.10, p=0.38) when depression and control groups were combined. Results were consistent with and without group and ICV as covariates.

### Imaging transcriptomics

The first PLS component explained 26.01% of the spatial variability in the strength of the association between CP volume and TSPO and did so above chance (r=0.51, p_spatial_ =0.006) (**Figure 6**). The list of genes ranked by the respective PLS1 weights can be found in “Supplementary data -[*PLS1_weights_CP_TSPO_Corr*]”. We found significant enrichment for six gene ontology (i.e. biological pathways terms among the top positively weighted genes), including “protein localization to endoplasmatic reticulum”, “leukocyte activation involved in inflammatory response”, “serotonin receptor signalling pathway”, “gamma-aminobutyric acid signalling pathway”, “neuroinflammatory response”, and “interleukin-1 response”. In the brain cell type enrichment analysis, we found enrichment among the top positively weighted genes mostly for genes from excitatory and inhibitory neurons, where the strongest positive enrichment hit was the subclass of excitatory neurons “Ex3e” (**Figure 6**) (Full statistics can be found in “Supplementary data -[*GSEA_PLS1_CP_TSPO_Corr*]”).

**Figure 6.**
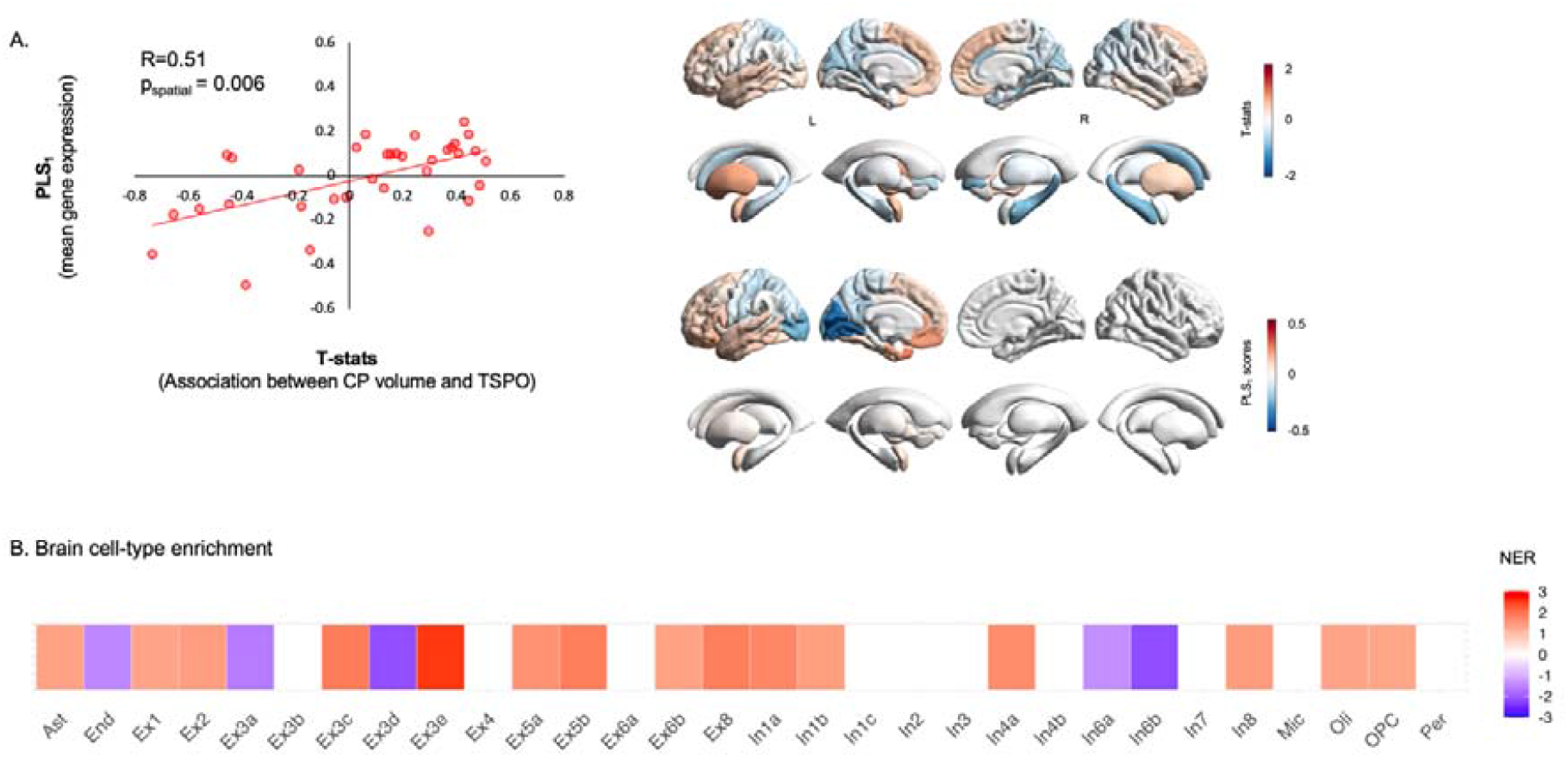
Imaging transcriptomics decoding of regional variation in the association between choroid plexus (CP) volume and TSPO. Panel A: Left - scatter plot depicting a significant positive correlation between PLS1 gene expression weights and the t-statistics quantification the association between CP volume and TSPO for each region of the left hemisphere. Right – the upper brain map depicts the cortical distribution of the t-statistics quantifying the association between CP volume and TSPO; the lower brain map depicts the cortical distribution of the weights of PLS_1_. Panel B: Table showing the results of the brain cell-type gene enrichment analysis. The colours depict normalized enrichment ratios; positive ratios indicate enrichment for genes of a specific cell class among those genes with the highest positive PLS_1_ weights; the reverse applies to negative enrichment ratios. White squares indicate cell classes where enrichment did not survive p_FDR_<0.05. NER – Normalized enrichment ratio; CP – Choroid plexus.

For the analyses on case-control differences in TSPO, PLS_1_ explained 33.64% of the spatial variability in TSPO differences and did so above chance (r=0.58, p_boot_=0.001) (Supplementary - Figure 1). In the gene set enrichment analysis, we did not find significant enrichment for any gene ontology. In the brain cell type enrichment analysis, we also did not find significant enrichment among positively weighted genes for genes from any of the cell types we tested (Full statistics can be found in Supplementary [*PLS1_weights_TSPO_Differences*] and [*GSEA_PLS1_TSPO_Differences*]).

## Discussion

We demonstrated for the first time the association between CP enlargement in depression and a reduction of brain barrier permeability reflected by blood-to-CSF radiotracer exchange parameters (SUVR). Moreover, we have also demonstrated that in the same patients CP volume is positively associated with increases in brain inflammation as measured by TSPO PET in key regions (ACC, PFC and Insular Cortex). There was no association between CP volume and peripheral inflammatory markers (CRP, IL6, and TNF-α) nor depression clinical scores (HDRS, Childhood Trauma score), but there was an association with Perceived Stress score. However, as this result was from exploratory analysis and did not survive multiple comparison testing correction, it should be taken with caution. Our results support the hypothesis that CP enlargement occurs in depressed cohorts, and this enlargement is associated with central inflammation.

The finding that CP volume is correlated with inflammation of the CNS but not with peripheral inflammation fits into the proposed model of the relationship between peripheral and central inflammation. We can speculate that CP volume may increase with time, likely together with the BBB dysfunction (Kealy et al., 2020) as a reaction to a heightened concentration of peripheral immune mediators. The lack of correlation between CP volume and CRP (which trends towards a positive association but does not achieve significance) is probably due to the CRP concentrations being an instantaneous measurement (Felger et al., 2018), while CP volume increase is likely the result of the integrated series of peripheral immune events across time (Marques and Sousa, 2015, Turner et al., 2014). Interestingly, CP volume increases were associated with reductions in plasma albumin which is a well established marker of chronic inflammation (Don and Kaysen, 2004). The enlargement of the CP volume in our result is associated with the reduction of the BCSFB permeability, which was identified as a reaction to the chronic inflammatory events in depressed subjects (Turkheimer et al., 2020). Similarly, the correlation between CP volume and brain inflammation may indicate that microglial activation is also the result of disturbed homeostasis across time (Enache et al., 2019). In our study, we did not see any elevation of CP [11C]PK11195 in depression, but we were able to replicate the association between CP volume and CP TSPO uptake as reported by in relapsing remitting multiple sclerosis patients (Ricigliano et al., 2021).

The spatial profile of the correlation pattern between CP volume and TSPO brain expression was positively associated with the neuronal distribution, twice stronger for excitatory neurons than for inhibitory ones (supplementary data). This result indicates that those brain areas metabolically more active are also those where inflammation is likely the result of reduced permeability of the barriers. These areas are known as “neotenic” as they contain a disproportionate density of synapses that have exceptionally high energy requirements and matching needs of efficient transport and clearance of metabolic substrates and products (Goyal et al., 2014, Vaishnavi et al., 2010). These results confirm an associative link between brain metabolism, brain barriers, permeability, and neuroinflammation. But further research is required to establish a causal link between these factors in MDD.

It is important to note that imaging data on the structure of CP do not explain the functional mechanism underlying such morphological modifications. Previous gene expression analysis from a cross-sectional post-mortem study has identified multiple differences in CP samples obtained from patients with depression, particularly in the downregulation of genes related to the “transforming growth factors-beta” (TGF-B) pathway, which are known to interact with the production of the extracellular matrix, suggesting changes in the cytoskeleton of CP epithelial cells in patients with MDD (Turner et al., 2014). Reduction of VEGF, a neurotrophic factor which is produced by the CP epithelial cells and responsible for angiogenesis and vascular fenestration permeability, were found in serum of suicide victims with MDD (Isung et al., 2012) and in plasma of MDD patients (Dome et al., 2009). To date, however, CP alterations remain poorly understood and multiple mechanisms are put forward that include CP ependymal cell proliferation (Barkho and Monuki, 2015), infiltration of CP by peripheral immune cells or edema (Johanson and Johanson, 2018).

Finally, it is important to note that the findings above do not seem to be specific to depression. Hence, this peripheral-to-central immunity mechanism seems to be generalizable to all pathologies where peripheral inflammatory states are observed, such as in schizophrenic cohorts (Lizano et al., 2019, Zhou et al., 2020). Recenlty, strong evidences from multiple sclerosis patients, a cuprizone diet-related demyelination mouse model and the experimental autoimmune encephalomyelitis model demonstrated a cross-dependency between neuroinflammation and choroid plexus functional, cellular and morphological characteristics which included volume enlargement, glia hyperactivity and the up-regulation of functional pathways primary related to neuroinflammation and cell-to-cell interactions (Fleischer et al., 2021). Taken together, these results further support the use of CP volume as a reliable, trans-diagnostic marker of neuroinflammation.

## Limitations

Some limitations have to be considered regarding our results. Manual CP segmentation is operator dependent but reanalysis of the CP volume with a second operator (JS) lead to an intraclass correlation coefficient ICC = 82% indicating robustness in our estimation. Automatic methods to measures CP volume exists but do not match well with manual extraction (Lizano et al., 2019) especially when applied to structural T1-weighted MRI data. The sample size in this study was small compared to recent big-data structural MRI studies in depression (N>1,000) (Schmaal et al., 2020). However, this is one of the largest samples ever published in depression that combines MRI with TSPO PET imaging. Future studies should investigate the use of automatic methods for CP segmentation in order to scale up these analyses to a larger sample size. Moreover, the collection of TSPO PET data with arterial input function to quantify tracer kinetic exchange though BCSFB would be more accurate than static SUVR and AUC parameters (Schubert et al., 2019) for measuring CSF dynamics.

## Conclusion

This study has identified for the first time in vivo a relationship between CP volumetric and brain inflammatory alterations in depression. The change on the CP morphology has been found in different neuroinflammation disorders, with support the role of CP enlargement in neuroinflammation. This study also provides further evidence to the model that reduced permeability of the BBB and BCSFB could alter brain homeostasis, becoming harmful if protracted by chronic inflammatory states. These findings do not seem to be specific to depression, but rather explain a more general mechanism of brain immune defence, in which CP plays a fundamental role.

## Supporting information

The list of genes ranked by the respective PLS1

## Data Availability

The datasets generated during and/or analysed during the current study are available from the corresponding author on reasonable request and upon approval from the research consortia.The dataset is part of "Biomarkers in Depression (BIODEP) study, NIMA consortium, https://www.neuroimmunology.org.uk/.

## Acknowledgements

The BIODEP study was sponsored by the Cambridgeshire and Peterborough NHS Foundation Trust and the University of Cambridge, and funded by a strategic award from the Wellcome Trust (104025) in partnership with Janssen, GlaxoSmithKline, Lundbeck and Pfizer. Recruitment of participants was supported by the National Institute of Health Research (NIHR) Clinical Research Network: Kent, Surrey and Sussex & Eastern. E Bullmore and CM Pariante are supported by a Senior Investigator award from the NIHR. Additional funding was provided by the National Institute for Health Research (NIHR) Biomedical Research Centre at South London and Maudsley NHS Foundation Trust and King’s College London, and by the NIHR Cambridge Biomedical Research Centre (Mental Health). Study data were collected and managed using REDCap electronic data capture tools hosted at the University of Cambridge (70). We would like to gratefully thank all study participants, research teams and laboratory staff, without whom this research would not have been possible. We thank and acknowledge all members of the NIMA Consortium at the time of data collection: Dominika Wlazly, Amber Dickinson, Andy Foster, Clare Knight, Claire Leckey, Paul Morgan, Angharad Morgan, Caroline O’Hagan, Samuel Touchard, Shahid Khan, Phil Murphy, Christine Parker, Jai Patel, Jill Richardson, Paul Acton, Nigel Austin, Anindya Bhattacharya, Nick Carruthers, Peter de Boer, Wayne Drevets, John Isaac, Declan Jones, John Kemp, Hartmuth Kolb, Jeff Nye, Gayle Wittenberg, Gareth Barker, Anna Bogdanova, Heidi Byrom, Diana Cash, Annamaria Cattaneo, Daniela Enache, Tony Gee, Caitlin Hastings, Melisa Kose, Giulia Lombardo, Nicole Mariani, Anna McLaughlin, Valeria Mondelli, Maria Nettis, Naghmeh Nikkheslat, Carmine Pariante, Karen Randall, Julia Schubert, Luca Sforzini, Hannah Sheridan, Camilla Simmons, Nisha Singh, Federico Turkheimer, Vicky Van Loo, Mattia Veronese, Marta Vicente Rodriguez, Toby Wood, Courtney Worrell, Zuzanna Zajkowska, Brian Campbell, Jan Egebjerg, Hans Eriksson, Francois Gastambide, Karen Husted Adams, Ross Jeggo, Thomas Moeller, Bob Nelson, Niels Plath, Christian Thomsen, Jan Torleif Pederson, Stevin Zorn, Catherine Deith, Scott Farmer, John McClean, Andrew McPherson, Nagore Penandes, Paul Scouller, Murray Sutherland, Mary Jane Attenburrow, Jithen Benjamin, Helen Jones, Fran Mada, Akintayo Oladejo, Katy Smith, Rita Balice-Gordon, Brendon Binneman, James Duerr, Terence Fullerton, Veeru Goli, Zoe Hughes, Justin Piro, Tarek Samad, Jonathan Sporn, Liz Hoskins, Charmaine Kohn, Lauren Wilcock, Franklin Aigbirhio, Junaid Bhatti, Ed Bullmore, Roido Manavaki, Sam Chamberlain, Marta Correia, Anna Crofts, Tim Fryer, Martin Graves, Alex Hatton, Manfred Kitzbichler, Mary-Ellen Lynall, Christina Maurice, Ciara O’Donnell, Linda Pointon, Peter St George Hyslop, Lorinda Turner, Petra Vertes, Barry Widmer, Guy Williams, Jonathan Cavanagh, Alison McColl, Robin Shaw, Erik Boddeke, Alison Baird, Stuart Clare, Phil Cowen, I-Shu (Dante) Huang, Sam Hurley, Simon Lovestone, Alejo Nevado-Holgado, Elena Ribe, Anviti Vyas, Laura Winchester, Madeleine Cleal, Diego Gomez-Nicola, Renzo Mancuso, Hugh Perry, Mara Cercignani, Charlotte Clarke, Alessandro Colasanti, Neil Harrison, Rosemary Murray, Jason O’Connor, Howard Mount.

